# Non-detection of emerging and re-emerging pathogens in wastewater surveillance to confirm absence of transmission risk: a case study of polio in New York

**DOI:** 10.1101/2023.08.24.23294561

**Authors:** David A Larsen, Dustin Hill, Yifan Zhu, Mohammed Alazawi, Dana Chatila, Christopher Dunham, Catherine Faruolo, Brandon Ferro, Alejandro Godinez, Brianna Hanson, Tabassum Insaf, Dan Lang, Dana Neigel, Milagros Neyra, Nicole Pulido, Max Wilder, Nan Yang, Brittany Kmush, Hyatt Green

## Abstract

Infectious disease surveillance systems, including wastewater surveillance, can alert communities to the threat of emerging pathogens. We need methods to infer understanding of transmission dynamics from non-detection. We estimate a sensitivity of detection of poliovirus in wastewater to inform the sensitivity of wastewater surveillance for poliovirus using both a clinical epidemiology and fecal shedding approach. We then apply freedom from disease to estimate the sensitivity of the wastewater surveillance network. Estimated sensitivity to detect a single poliovirus infection was low, <11% at most wastewater treatment plants and <3% in most counties. However, the maximum threshold for the number of infections when polio is not detected in wastewater was much lower. Prospective wastewater surveillance can confirm the absence of a polio threat and be escalated in the case of poliovirus detection. These methods can be applied to any emerging or re-emerging pathogen, and improve understanding from wastewater surveillance.

## Introduction

Increasing vaccine hesitancy^1^ and pandemic-disrupted childhood vaccine schedules^2^ raise the potential for polio outbreaks in countries where polio had previously been eliminated. For example in 2022, London, England saw sustained transmission of polio (but no paralytic cases)^3^ and New York State had a paralytic polio case and detection of poliovirus in wastewater also in 2022.^4,5^ Although completely effective at preventing paralysis from a polio infection, the inactivated polio vaccine administered in the US and other wealthy countries is not completely effective at preventing onward transmission.^6^ Further, county- and state-level vaccine coverages mask gaps in community-level vaccine coverages, with pockets of communities with very low vaccine uptake.^7^

In the United States, the transmission of poliovirus is currently monitored through acute flaccid paralysis (AFP) surveillance and follow-up testing for poliovirus. However, AFP is estimated to occur only once in 200 wildtype polio infections and once in 2,000 infections of vaccine-derived poliovirus (VDPV).^8^ Moreover, with high polio vaccination coverage the vast majority of infections are asymptomatic^9^ suggesting that poliovirus could circulate without AFP surveillance providing any indication.

Testing for poliovirus in wastewater complements AFP surveillance across the globe,^10^ often to good effect. For example, in Pakistan, polio circulation was detected via wastewater an average of four months before AFP surveillance.^11^ And in Israel, detection of poliovirus in wastewater prompted a vaccine campaign that was able to prevent any paralytic cases of polio.^12^ Despite the utility of wastewater surveillance for polio and its widespread implementation, its sensitivity is not well defined. Previously, the sensitivity of wastewater surveillance to detect polio has been estimated to be 35-50% in Afghanistan and Pakistan depending on the sampling site and immunization activities.^8^ These estimates are helpful, but are only derived from a single context. Accurate estimates of sensitivity are essential for estimating the probability of not only detecting an emerging pathogen, but also that the community is free from transmission upon consecutive non-detections following freedom from disease principles.^13,14^

In July of 2022 a local case of VDPV was confirmed in a patient with AFP in Rockland County, New York.^4^ Immediately in coordination with the CDC, New York State’s wastewater surveillance network began testing for polio in the wastewater throughout Rockland County, surrounding counties, New York City, and Long Island.^5^ Widespread detection of poliovirus in the wastewater of Rockland County and five surrounding jurisdictions led New York State to declare a disaster emergency.^15^ To aid interpretation of the non-detection of poliovirus in wastewater, we estimated the sensitivity of wastewater in detecting circulating poliovirus within each county and individual sewersheds using two different approaches: one based on spatiotemporal coverage of a represented population and another based on mass balance and fecal shedding. Based partially on these estimates, we outline a wastewater surveillance plan considering polio vaccine coverage to ensure both the elimination of polio transmission and provide early indication of any polio re-emergence.

## Methods

### Estimating sensitivity of wastewater surveillance of poliovirus

The sensitivity of an infectious disease surveillance system to detect emerging threats such as polio is a product of the population coverage, the temporal coverage, and the sensitivity of detection.^16^ For wastewater surveillance, the sensitivity of detection is determined by the limit of detection of the pathogen in wastewater, which is typically driven by the population served (dilution) and the amount of genetic material shed into wastewater. We used equation 1 to estimate the sensitivity of wastewater surveillance for poliovirus. We defined population coverage (C_p_) as the proportion of the population connected to the wastewater surveillance network. For estimates at the treatment plant level we assumed 100%. For estimates at the county level we use the calculated proportion of the population connected to the wastewater surveillance network from our previous work.^17^ We relate temporal coverage (C_t_) to the length of poliovirus fecal shedding. Polio infections on average shed virus in feces for 3-4 weeks (midpoint of 25 days).^18^ We divided 25 days by the number of days between consecutive wastewater sampling events. For sampling intervals smaller than 25 days we assigned C_t_ a value of 100%. We estimated the sensitivity of detection (Se_d_) in two ways. First, we analyzed the New York State polio outbreak of 2022.^4^ Second, we used modeled estimates of viral copy detection per wastewater treatment plant flow.

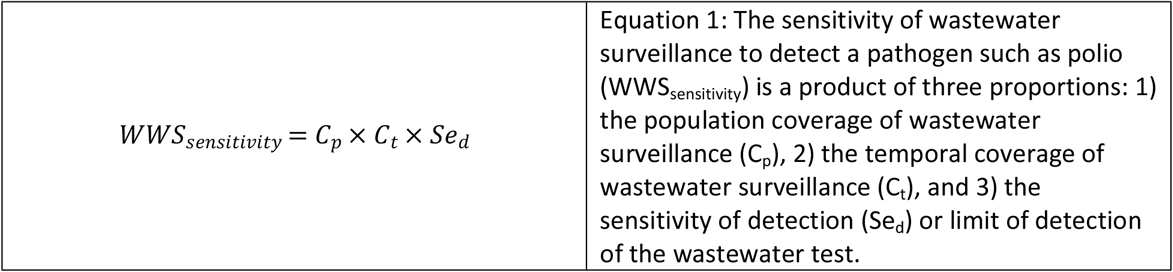

**The New York State polio outbreak of 2022** was identified when a young adult presented to the hospital with AFP caused by vaccine-derived poliovirus.^4^ No other paralysis cases have yet been observed (as of June 2023), despite numerous detections of poliovirus DNA in wastewater in multiple communities. From the literature we assumed that one paralysis case of vaccine-derived poliovirus occurs for every 2,000 infections.^8^ Equation 2 shows our approach to estimating the sensitivity of detection from the New York State polio outbreak. We first simulated the total number of polio infections among unvaccinated individuals with 10,000 iterations (n) of a binomial distribution (B) using a probability of 1 paralytic case per 2,000 infections (p). From this number of infections we calculated a prevalence of polio among unvaccinated individuals (prev_unvaccinated_). Assuming equal mixing among unvaccinated and vaccinated populations, we applied the prevalence of polio among unvaccinated individuals to the population of vaccinated individuals and estimated the number of polio infections among vaccinated individuals. We then distributed the total number of infections across the sewersheds where poliovirus was detected proportional to the population of the sewershed. Lastly, the population size required for 1 successful detection in the sewershed was divided by population to get the detection sensitivity.

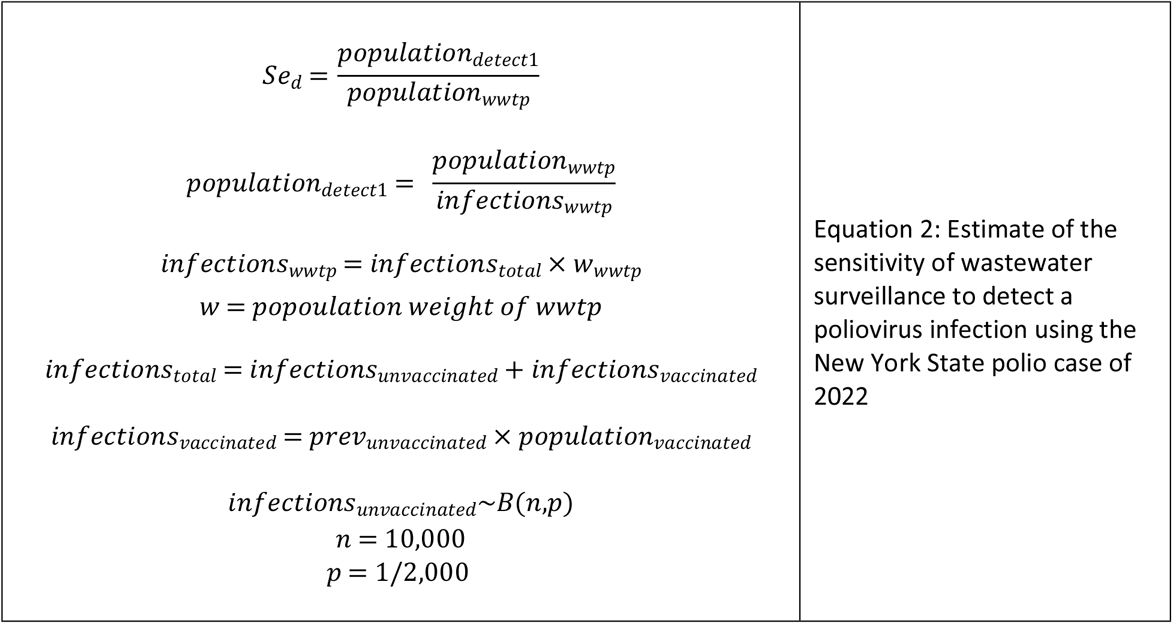

**Fecal shedding studies of poliovirus** are limited with fecal shedding studies reporting duration and temporality of shedding but not quantity.^18^ Still, Berchenko et al. estimated that wastewater surveillance was able to identify one poliovirus shedder per 400,000 liters of sewage following an oral polio vaccine campaign in Israel.^19^ We can then estimate the sensitivity of wastewater surveillance to identify poliovirus circulation as a function of a wastewater treatment plant’s daily flow using Berchenko et al.’s estimate (Equation 3). We applied the mean daily flow to estimates of sensitivity for each treatment plant. For any treatment plant without mean daily flow we applied an estimated mean ratio of daily flow to permitted discharge capacity of those treatment plants with data to those treatment plants without data.

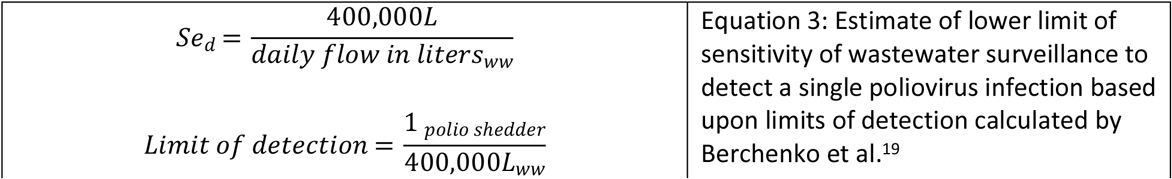

### Understanding non-detection of poliovirus

With the estimated sensitivity of the wastewater surveillance network to detect poliovirus from equation 1, we calculated the probability that a community was free from poliovirus circulation (freedom from disease) using equation 4. From the two estimated sensitivities of detection (Se_d_ in equations 2 and 3) we applied whichever was lower. We estimated this probability for three consecutive non-detections of poliovirus in wastewater for all treatment plants in the state network and for each NY state county. Reversing equation 1, a sensitivity of 63% is required to obtain 95% confidence in zero infections with three consecutive non-detections. We estimated an upper limit of the number of poliovirus infections present with three consecutive non-detections of poliovirus in wastewater for all treatment plants in the state network and for each NY state county using equation 5.

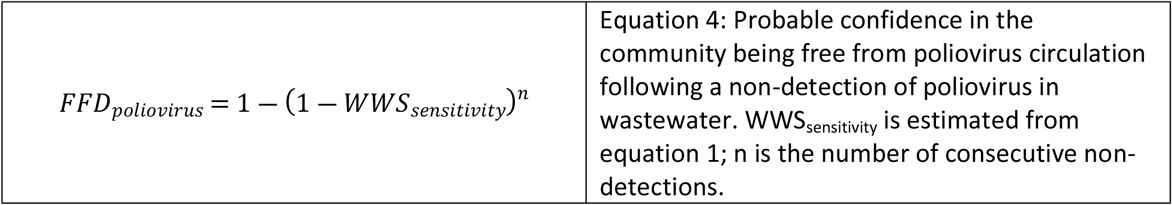

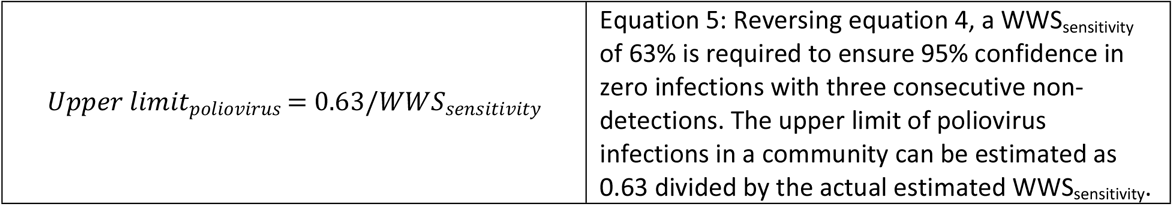

### Spatial comparison of vaccination rates

We compared zip code level polio vaccination rates provided by New York State Department of Health to municipal wastewater treatment plant (WWTP) catchment areas in NY with permitted discharge capacity of at least 1 million gallons per day (mgd). Zip codes with vaccination rates of less than 70% that intersected with sewersheds were flagged as being areas that were both extra vulnerable to polio transmission and also within the wastewater surveillance network. Sewersheds that intersected with these zip codes were then classified for whether they would be routine participants in the state’s wastewater surveillance network (i.e., permitted discharge capacity above 1 mgd) or if they would not be routinely tested but could be enrolled for testing under special circumstances. Zip codes that did not intersect with any sewersheds were also noted as potential blind spots for the use of wastewater to test for polio presence.

We also explored other potential risk factors for polio circulation including social vulnerability. We estimated the Centers for Disease Control’s (CDC) Social Vulnerability Index (SVI)^20^ for each sewershed. The SVI uses U.S. Census data to identify if a community might have greater vulnerability to external stressors that might make the area at higher risk for negative outcomes of natural disasters or disease outbreaks. SVI values range from 0 to 1 with higher indexes being more vulnerable. We calculate the mean SVI for each sewershed from the NY census tracts that intersected with the sewershed. We then assessed correlation between the SVI and polio vaccine coverage using a Pearson correlation test. All spatial analyses were conducted in R programming software^21^ using the package “sf”.^22^

## Results

### Estimating sensitivity of wastewater surveillance of poliovirus

From the New York State polio outbreak we estimated 1,401 unvaccinated individuals infected with polio (95% CI = 1,359-1,439) in the counties with polio found in the wastewater. This represents a prevalence of 0.13% (95% CI = 0.12 – 0.13%) among the 1,115,881 unvaccinated individuals across the study area. Assuming the same prevalence among the 4,329,977 vaccinated individuals across the study area we estimated 5,436 vaccinated individuals infected with polio (95% CI = 5,273 – 5,584). After distributing all 6,837 estimated polio infections (95% CI = 6,632 – 7,023) across wastewater treatment plants proportional to the population served by the wastewater treatment plant we estimate that wastewater surveillance was consistently able to detect one polio infection among 472 people connected to the sewer system (95% CI = 460 – 487). This resulted in a sensitivity of detection of a single polio infection ranging from 0.04% in the largest treatment plant in the study area to 12.2% in the smallest (Table 1).

**Table 1:** Estimates of the probability of having at least one paralytic case among unvaccinated infections and lower limits of sensitivity of detection of poliovirus in wastewater based upon a single paralytic polio case in Rockland County, NY and subsequent detections of poliovirus in wastewater in surrounding counties using the New York State polio outbreak.

| County | Treatment plant | WWTP capacity (millions of liters) | Population served | Paralytic polio cases | Polio vaccine coverage | Estimated polio infections (95% CI) | Approach #1 estimated sensitivity of detection (95% CI) | Approach #2 estimated sensitivity of detection (95% CI) | Approach #3 estimated sensitivity of detection (95% CI) |
| --- | --- | --- | --- | --- | --- | --- | --- | --- | --- |
| Kings (Brooklyn) | Coney Island | 416 | 711,000 | 0 | 85% | 1505 (1459 – 1545) | 0.07% (0.06 – 0.07%) | 0.13% (0.07 – 0.22%) |  |
| Kings (Brooklyn) | Newtown Creek | 1,173 | 1,197,000 | 0 | 87% | 2533 (2457 – 2602) | 0.04% (0.04 – 0.04%) | 0.05% (0.03 – 0.09%) |  |
| Kings (Brooklyn) | Owls Head | 454 | 825,000 | 0 | 86% | 1746 (1693 – 1793) | 0.06% (0.06 – 0.06%) | 0.11% (0.06 – 0.18%) |  |
| Nassau | Port Washington | 15 | 21,000 | 0 | 66% | 44 (43 – 46) | 2.3% (2.2 – 2.3%) | 3.87% (2.29 – 6.52%) |  |
| Orange | Harriman | 23 | 43,000 | 0 | 53% | 91 (88 – 93) | 1.1% (1.1 – 1.1%) | 2.24% (1.32 – 3.76%) |  |
| Richmond (Staten Island) | Port Richmond | 227 | 211,000 | 0 | 86% | 447 (433 – 459) | 0.2% (0.2 – 0.2%) | 0.38% (0.22 – 0.64%) |  |
| Rockland | Rockland County #1 | 109 | 207,000 | 1 | 76% | 438 (425 – 450) | 0.2% (0.2 – 0.2%) | 0.48% (0.29 – 0.81%) |  |
| Rockland | Western Ramapo | 5.7 | 4,000 | 0 | 66% | 9 (8 – 9) | 11.8% (11.5 – 12.2%) | 8.50% (5.04 – 14.32%) |  |
| Sullivan | Monticello | 11.7 | 7,000 | 0 | 73% | 15 (14 – 15) | 6.8% (6.6 – 7.0%) | 4.92% (2.92 – 8.30%) |  |
| Sullivan | South Fallsburg | 10 | 5,000 | 0 | 57% | 11 (10 – 11) | 9.5% (9.2 – 9.7%) | 4.83% (2.86 – 8.13%) |  |

Using Berchenko et al’s estimate of wastewater surveillance being able to detect one polio infection per 400,000 liters of flow (95% CI: 231,000-656,000),^19^ we find the sensitivity of detection to range from 0.05% in the largest treatment plants in the study area to 8.5% in the smallest (Table 1).

### Understanding non-detection of poliovirus

Figure 1A shows estimates of sensitivity to detect a single poliovirus infection at the wastewater treatment plant level across New York State, using whichever estimated sensitivity of detection (Se_d_) is higher from the two different approaches. Three consecutive non-detections of poliovirus provide confidence in zero poliovirus infections in the community ranging from 0.2% to 99% (Figure 1B) and 95% confidence that the number of infections within a community range from 0 to fewer than 1,079 (Figure 1C).

**Figure 1:**
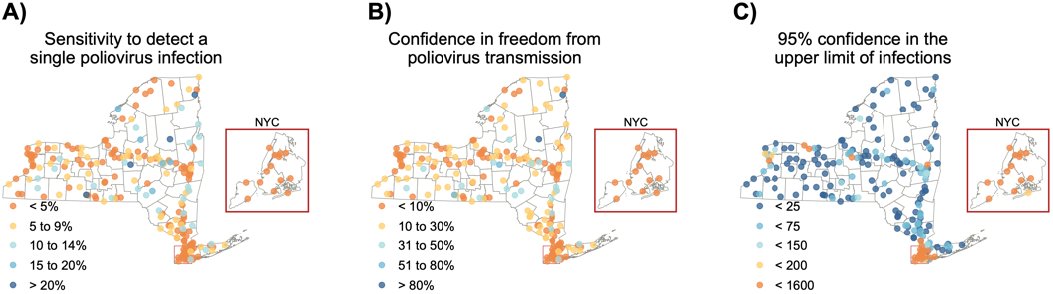
The sensitivity of wastewater treatment plants in New York State to a) detect a single poliovirus infection, b) provide confidence in the freedom from poliovirus transmission with three consecutive non-detections, and c) provide 95% confidence in the upper limit of the number of poliovirus infections with three consecutive non-detections.

Once applying county-level population coverage, we find the sensitivity of wastewater surveillance to detect a single poliovirus infection to range from 0.5% in New York County (Manhattan) to 10.7% in Clinton County (Figure 2A). Three consecutive non-detections of poliovirus provide confidence in zero poliovirus infections in the county ranging from 0.1% to 28.7% (Figure 2B) and 95% confidence that the number of infections within a county range from fewer than 6 to fewer than 1,570 (Figure 2C).

**Figure 2:**
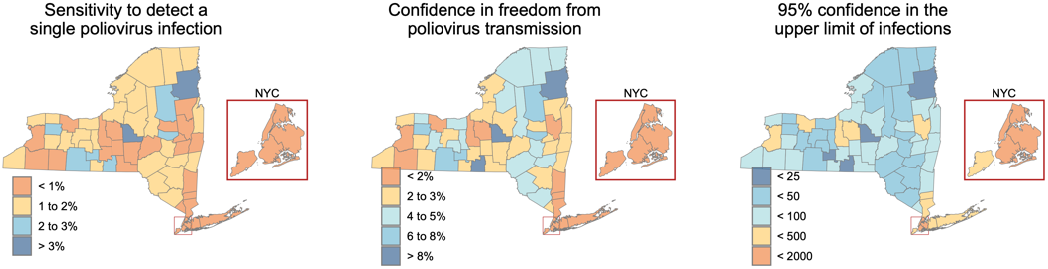
The sensitivity of New York State’s wastewater surveillance to identify at the county level a) a single poliovirus infection, b) confidence in the freedom from poliovirus transmission with three consecutive non-detections, and c) an upper limit of the number of poliovirus infections given three consecutive non-detections.

### Spatial comparison of vaccination rates

Sewersheds with higher social vulnerability had lower vaccination rates (Pearson correlation coefficient of -0.2298, p value < 0.01). The majority of sewersheds in New York (n=459, 76.6%) have vaccination rates above 70% and lower vaccination coverage sewersheds (n=140, 23.4%) ranged in vulnerability between 0.1 and 0.85 with a median SVI of 0.51. Sewersheds with an average vaccination rate greater than 70% ranged between an SVI of 0 and 0.8 with a median SVI of 0.45.

An estimated 1.76 million New Yorkers reside in zip codes with poliovirus vaccination coverage < 70%. The majority of these New Yorkers (81%) are covered by the wastewater surveillance network (Table 2). A further 11% of these New Yorkers live in communities connected to sewer, but their wastewater treatment plants are not currently enrolled . (The threshold for inclusion in the state’s wastewater surveillance network is a treatment plant permitted to discharge at least 1 million gallons per day). Eight percent of New Yorkers living in zip codes with poliovirus vaccination coverage < 70% are not connected to any public sewer system.

**Table n:** Number of zip codes across NY with vaccination rates below 70% and whether they are within a sewer system that can be monitored using wastewater surveillance.

|  |  |  |  |
| --- | --- | --- | --- |
| Status of zip code | Number of zip codes with vaccination rates <70% | Total population of the zip codes | Percent of total |
| Zip code included in network and eligible for ongoing surveillance | 87 | 1,420,000 | 81 |
| Zip code intersects with a small sewershed and could be included in potential scale-up of surveillance | 67 | 190,000 | 11 |
| Zip code not in a sewershed | 115 | 150,000 | 8 |
| Total | 269 | 1,760,000 | 100 |

## Discussion

We find that the sensitivity of wastewater surveillance to detect any circulating poliovirus is highly dependent upon the size of the population contributing to the wastewater sample and unique to each sewershed. In general, when using the input parameters herein the sensitivity of a single test of wastewater collected from a wastewater treatment plant to detect poliovirus is low, < 10% throughout much of New York State. This issue is compounded by gaps in population coverage of wastewater surveillance when estimating county-level sensitivity, < 2% sensitivity throughout much of New York State. Three consecutive non-detections of poliovirus are slightly better rising at treatment plants throughout the state and but still < 5% sensitivity at the county level. Despite the low sensitivities observed, non-detection of poliovirus in wastewater can still provide maximum thresholds of the number of poliovirus infections within the sewer catchment or the county.

These estimates of sensitivity are primarily driven by the sensitivity of detection, which we consider in these analyses to be highly conservative. For example, we estimate a sensitivity of detection of one shedder per 472 population, much lower than the one shedder per 10,000 population that Hovi et al. estimated following flush experiments.^23^ If using Hovi et al’s one shedder per 10,000 population the confidence in no transmission is greatly increased, above 50% throughout much of New York State. We elected not to include results using Hovi et al’s estimated sensitivity of detection due to the huge difference between that method and the methods estimated from Berchenko and our crude modeling. The low sensitivity in these results further align with those by O’Reilly et al., who also found < 10% sensitivity in the surveillance system to identify polio transmission when the number of shedders is minimal.^23^

We consider these estimates to be conservative for a number of reasons. First, we rely heavily on Berchanko et al.^19^ in the absence of fecal shedding data. Fecal shedding studies that estimate the number of viral copies per gram of feces excreted among polio-infected individuals would allow for a more robust approach. Second, we presume equal mixing and equal onward polio transmission of vaccinated and unvaccinated populations. Polio vaccines are extremely effective at eliminating the risk of paralysis, however only oral polio vaccine is considered a transmission-interrupting vaccine. The inactivated polio vaccine that is administered throughout New York does not produce sterilizing immunity – vaccinated individuals can still contract and transmit polio albeit to a lesser degree than unvaccinated individuals.^24^ The exclusion of the effect of vaccines from estimates on the number of infected individuals likely overestimates the number of polio infections throughout New York State in Table 1. This would likely reduce our estimates of sensitivity to detect any poliovirus circulating. It is unlikely that poliovirus is widespread and circulating everywhere among individuals vaccinated with inactivated polio vaccine,^8^ otherwise we would see much more paralytic polio among unvaccinated individuals. To date (August 2023) in the Rockland County outbreak we have only seen a single paralytic case of polio, and this is the first polio case detected in New York since 2013. Furthermore, public health is primarily concerned with polio infections among vulnerable (unvaccinated) individuals. A small outbreak among vaccinated individuals that never reaches susceptible unvaccinated individuals is of minimal concern.

The most vulnerable communities to polio outbreaks (zip codes with < 70% vaccine coverage) are largely connected to the state’s wastewater surveillance network that we have established. In these communities 81% of residents are wholly encompassed within the network, and a further 11% of residents intersect in some way with smaller treatment plant catchments. It is likely that any polio outbreak in low vaccination communities would first be identified in wastewater, as has been observed elsewhere.^11,12^

We can apply these results to inform testing frequency in a statewide wastewater surveillance network (Figure 3). The cost of wastewater surveillance is primarily driven by the number of samples (the spatial scale) and frequency of sampling. The system should more often test communities vulnerable to outbreak including communities with lower vaccine coverage, larger cities, and globally connected communities. We suggest a baseline prospective surveillance sampling frequency of once every two weeks. If poliovirus is detected in wastewater, we recommend scaling up the sampling frequency to weekly and expanding the spatial coverage in the county with the detection and connected communities. Three consecutive non-detections prompt a reduction in temporal frequency, reducing to once every two weeks (scaling back surveillance). Once elimination is confirmed again (zero poliovirus found in consecutive samples in any area) then baseline prospective surveillance can resume. This is just one way of operationalizing wastewater surveillance system with guidance on where and how often to test wastewater.

**Figure 3:**
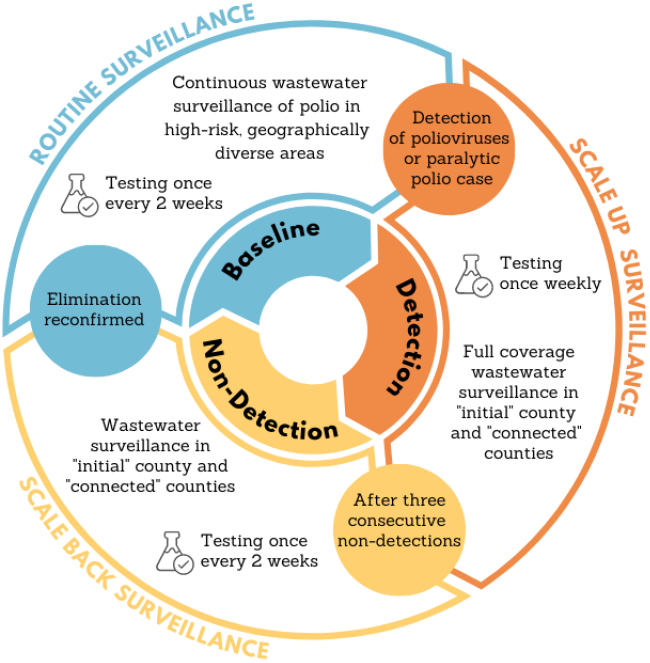
Conceptual framework to guide the frequency and scale of testing wastewater for poliovirus. Baseline prospective surveillance regularly tests wastewater from communities vulnerable to polio outbreaks. Detection prompts a scale-up in surveillance, increasing both the spatial coverage and temporal frequency. Consecutive non-detections prompt scaling back in surveillance, reducing the temporal frequency. Once elimination is confirmed the system returns to baseline prospective surveillance.

The methods we outline here can be used to estimate the sensitivity of wastewater surveillance for any pathogen.^25^ Required inputs are fecal/urinary shedding rates and/or case data. In the absence of incident polio cases without paralysis, we used an upper limit of the number of infections thought to occur before a paralytic case is observed. This approach was not needed for our estimates of the sensitivity of wastewater surveillance to detect COVID-19 infections, where we used reported case data,^13^ nor would it be needed for any infectious disease where case data more accurately reflect the number of infected individuals. As wastewater surveillance becomes more routine and established,^26^ the methods outlined here can inform public health understanding from non-detections. We expect these methods to be most valuable in the context of emerging and re-emerging diseases, or disease elimination situations.

## Data Availability

We can include most of the data as a supplemental file. The Zip-code level polio vaccination rates cannot be included due to privacy concerns.

